# Physical activity and favourable adiposity genetic liability reduce the risk of hypertension among high body mass individuals

**DOI:** 10.1101/2024.12.18.24319295

**Authors:** Chukwueloka Hezekiah, Raha Pazoki

**Affiliations:** Cardiovascular and Metabolic Research Group, Division of Biomedical Sciences, Department of Life Sciences, College of Health, Medicine and Life Sciences, Brunel University London, UB8 3PH, United Kingdom; Department of Mental Health, Faculty of Health, Science, Social Care and Education, Kingston University, Kingston Hill, Kingston Upon Thames, Surrey, KT2 7LB, United Kingdom; Department of Epidemiology and Biostatistics, School of Public Health, St Mary’s campus, Norfolk Place, Imperial College London, London W2 1PG, United Kingdom

**Keywords:** Hypertension, favourable adiposity, genetic liability, physical activity, body mass

## Abstract

**Background and Purpose:** Hypertension is a global health issue, and the risk factors include genetics, physical inactivity, and excess body fat (adiposity). Genetic predisposition to adiposity generally increases risk of hypertension. Several genetic variants have been identified to increase adiposity but unexpectedly reduce hypertension (favourable adiposity genes). Here, we tested the effect of these genetic variants on risk of hypertension in European ancestry participants under various scenarios of physical activity and body mass index.

**Methods:** Favourable adiposity genetic liability was estimated using previously identified genetic variants and their effect sizes. The study analysed data from 230,027 unrelated participants in the UK Biobank. Logistic regression was used to examine the association between this genetic liability and hypertension (defined as systolic blood pressure ≥ 140 mmHg, diastolic blood pressure ≥ 90 mmHg, or the use of anti-hypertensive medications). The analyses were conducted separately based on physical activity status (physically active and inactive) within low and high body mass groups.

**Results:** Individuals with high body mass, could reduce their risk of hypertension by up to 16% depending on the favourable adiposity genetic liability and physical activity status (P _adjusted_ = 1.32 x10^-8^). In high body mass individuals, physical activity alone contributes to 6-9% reduction in risk of hypertension.

**Conclusion:** The study provides evidence that the protective effect of favourable adiposity on hypertension risk varies according to body mass composition and physical activity status.

## Introduction

Hypertension is a significant global health concern affecting about 1.3 billion adults globally ^1^. Several risk factors for hypertension exist, including obesity indicated by high body mass ^2^ as well as adiposity indicated by excess body fat ^3^. A large body of evidence exist emphasizing that excess body mass ^4–6^ and total body fat mass percentage ^3^ increased risk of hypertension.

For over a decade, genome-wide association studies (GWAS) in the general population have identified common alleles associated with both obesity ^7,8^ and adiposity ^9^. The genetic liability that is driven by these genetic variants increase the risk of hypertension ^10,11^. Recently, A total of 36 genetic adiposity variants, associated with body fat percent, have been identified as favourable adiposity variants ^12^. Favourable adiposity phenotype is defined as carrying a low risk of cardiometabolic disorders while carrying a large body fat percent ^12^. Accumulating evidence from genetic studies suggests that individuals with favourable adiposity genes have lower risk of hypertension than individuals with unfavourable adiposity genes ^12,13^. For example, Yaghootkar and colleagues ^14^ reported an association between favourable adiposity alleles and a higher fat mass, a lower systolic and diastolic blood pressure, and a lower risk of hypertension ^14^. Individuals carrying favourable adiposity variants exhibit a healthy metabolic profile, reducing their risk of developing several conditions, including hypertension ^13,14^. Ahmed and colleagues found that the favourable adiposity genetic liability is associated with a healthy metabolic profile and a low systolic and diastolic blood pressure in various ethnic groups including Europeans ^13^. These findings confirm the beneficial effect of favourable adiposity alleles in reducing the risk of hypertension.

It is known that lifestyle factors such as regular physical activity can help reduce adiposity and mitigate the risk of hypertension ^11,15^. In our previous work, we observed that genetic underpinning of body mass index (BMI) increases the risk of hypertension and physical activity offsets this risk. ^11^. Here, we aim to test the effect of favourable adiposity genetic liability on hypertension in presence and absence of physical activity and for various subgroups of BMI within European ancestry participants of the UK Biobank (UKB). The research question is that whether physical activity offsets the effect of favourable adiposity on hypertension and what the pattern of association is among individuals with high and low BMI. Such research is crucial to provide more insight into personalised lifestyle choices, enhance effectiveness of treatments, leading to better management of hypertension and overall health outcomes.

## Methods

### Study Population

The UKB is a major prospective cohort study that started in 2006 to allow detailed investigations of genetic and non-genetic determinants of disease. Between 2006 and 2010 (the initial assessment), more than 500,000 individuals of mainly European ancestry aged 40 to 69 were recruited by the UKB ^16^. During the initial assessment in the UKB, a wide range of phenotypic information was gathered following informed consent. This data was collected through interviews and touch-screen questionnaires, including socio-demographic, lifestyle and health-related information. Participants also underwent physical and anthropometric measurements during this assessment. The participants provided saliva, urine, and blood samples for proteomic, genetic, and metabolomic analyses ^17^.

### Ethical Approval

Our study was conducted in accordance with the principles outlined in the declaration of Helsinki ^18^. The Northwest Multi-Centre Research Ethics Committee (11/NW/0382) granted the UKB ethics approval as a research tissue bank approval. Participants gave their informed consent, and the UKB handled ethical approval.

Using approved data request application ID 60549, data for this study was obtained from the UKB. The College of Health, Medicine, and Life Sciences Research Ethics Committee at Brunel University London provided ethical approval for the current analysis to work on secondary data from the UKB (reference 27684-LR-Jan/2021-29901 1).

### Genotyping and Imputation

The UKB did the genotyping and imputation centrally. Comprehensive techniques used by the UKB are provided elsewhere ^19–21^. In summary, the participants’ blood samples were collected at the assessment centre, and the DNA was extracted and genotyped by the UKB using the UKB Axiom Array. Three reference panels were utilised for genotype imputation: the Haplotype Reference Consortium, UK10K, and 1000 Genomes phase 3. IMPUTE4 programme was used to carry out the imputation. The UKB calculated the kinship coefficients and genetic principal components centrally. These helped them identify related individuals and account for population stratification ^19^.

### Sample for analysis and Exclusion Criteria

The current study was performed using a subset of unrelated European ancestry individuals from the UKB. Genetic data were available for 488,377 individuals, and after merging genetic and phenotype data, 486,976 individuals remained for analysis. The current study applied several exclusion criteria. In summary, we excluded participants who withdrew consent (n=321), first- or second-degree relatives (n=26,102), participants of non-European ancestry (n=27,121), sex mismatch (n=328), pregnant women or women unsure of pregnancy status at baseline (n=513), did not declare their smoking status (n=221). Furthermore, we also excluded participants with missing data in their pack-years of smoking (n=66,062), current or previous smokers for whom zero pack-years of smoking was calculated (n=1049), unsure about their dietary intake of fish, meat, fruits or vegetables (n=12,919) and participants on cholesterol-lowering medication (n=60,090). In addition, participants who did not declare drinking status (n=160), participants whose self-reported ancestry did not match their genetic ancestry (n=44) and individuals with missing data in the main study covariates (n=62,340) were also excluded. A total of 230,027 participants remained for analysis (**Supplementary** Figure 1).

### Blood Pressure and Definition of Hypertension

Blood pressure was measured centrally by the UKB. The details of the blood pressure phenotype have been described previously ^11^. In brief, systolic blood pressure (SBP) and diastolic blood pressure (DBP) were measured twice at the UKB assessment centre using an automated sphygmomanometer (Omron HEM 7015-T) or a manual sphygmomanometer where an automated blood pressure reading could not be taken. This study used the mean of these readings obtained using the same device (automated or manual sphygmomanometer) to calculate the average blood pressure (SBP and DBP). Where blood pressure values obtained from different devices existed, we calculated average blood pressure using values from those devices. For participants on blood pressure-lowering medication, we added 15 mmHg to the SBP and 10 mmHg to the DBP ^22^ (n=89,524). We defined hypertension as stage 2 according to the American Heart Association guidelines ^23^ , i.e. SBP ≥ 140 or DBP ≥ 90 mmHg. We additionally defined the use of blood pressure-lowering medications as hypertension.

### Physical Activity Status

The details of the physical activity phenotype have been described previously ^11^. In summary, the UKB used a modified version of the short International Physical Activity Questionnaire (IPAQ) to measure physical activity ^24^ using a touch screen questionnaire. They collected data on the frequency, intensity and duration of walking, moderate and vigorous physical activity. Cassidy and colleagues ^25^ calculated the MET minutes per week of physical activity using the IPAQ data processing guidelines ^26^, and the data was returned to the UKB. The current study used this returned data from Cassidy and colleagues ^25^ to categorise participants into physically active (moderate activity ≥150 minutes per week or vigorous activity ≥75 MET minutes per week or summed MET minutes per week for all activities ≥ 600). We categorised participants who did not meet these criteria into a physically inactive group.

### Obesity Status

UKB determined the BMI values during the initial assessment centre visit by dividing weight in kilograms (measured using the Tanita BC-418 MA body composition analyser) by height in metres (measured using a Seca 202 device). If either of these values were absent, the UKB omitted the BMI value for those participants ^16^. The current study used the BMI values to define the participants’ obesity status. We classified BMI (Kg/m2) using the World Health Organisation’s categories: underweight (<18.5), normal weight (≥18.5 to <25), overweight (≥25 to <30), and obese (≥30) ^27^. In the present study, we categorised participants using the BMI values into (1) high body mass (overweight and obese) and (2) low body mass (normal weight and underweight).

### Assessment of covariates

The UKB used a self-reported questionnaire to assess participants’ dietary intake. Using touchscreen multiple-choice questions, participants selected their daily food consumption (vegetable, fruit, oily fish and meat intake). Smoking status in the UKB was determined through self-reported questions and categorised into current, past, and never smoking. Pack-years of smoking were available for the sample. The Pack-years of smoking were calculated based on the number of cigarettes smoked per day, divided by the average pack-size of twenty, and multiplied by the number of years smoking ^28^. This current study defined smoking based on the values from smoking status and pack-years of smoking. We defined smokers as current or previous smokers with any value in pack-years. Never smokers were those who reported never smoking and had zero values in their pack-years of smoking. We also included alcohol drinking status (current, past and non-drinkers) ^29^, and low-density lipoprotein (LDL) cholesterol as additional covariates in the analysis. Detailed quality control and sample processing information for the UKB biomarker data has been published previously ^30^.

### Favourable adiposity genetic liability

We estimated favourable adiposity genetic liability based on a list of previously published single nucleotide polymorphisms (SNPs) associated with favourable adiposity from Martin and colleagues ^12^. We used the effect estimates of these SNPs on favourable adiposity as their weight in calculation of the genetic liability. Using LDlink ^31^, we assessed the linkage disequilibrium (LD) among the SNPs. As part of the SNP selection process in this study, we considered only SNPs that met the GWAS significance threshold of P < 5 x 10^-8^, and SNPs with Minor allele frequency >0.01. We defined SNP pairs with R^2^ > 0.1 as correlated. Therefore, only SNPs that did not exhibit dependence on another SNP based on the LD parameter of R^2^ < 0.1 were considered. The final number of SNPs available for estimating the genetic liability for favourable adiposity was 35 (**Supplementary Table 1**). One SNP (rs555162510) that was not present in the 1000 Genomes Project data was removed. Using Plink v2.0 ^32,33^ to calculate genetic liabilities, we multiplied the number of risk alleles each UKB participant carries on the favourable adiposity SNPs by their effect estimate. The products were summed up to generate the genetic liability which was then standardised using the *base* package in R. The association results were therefore presented per standard deviation increase in the genetic liability of favourable adiposity.

### Statistical Analysis

We conducted a cluster analysis using the K-means algorithm to determine the participants’ genetic distance ^34^. A parameter indicating the number of clusters (K) is needed for the K-means algorithm ^35^. The number of categories within the UKB self-reported phenotypic variable, ethnic background, led to assigning a ’K’ value of seven (**Supplementary Table 2**). We compared the genetically derived clusters and self-reported ancestries to determine which participants’ self-reported ancestry did not match the genetically derived ancestry.

We compared the prevalence of hypertension within different categories of favourable adiposity genetic liability (moderate and high) against a reference group consisting of participants with low genetic liability. Using logistic regression, we examined the association between genetic liability and risk of hypertension across high and low body mass groups. We additionally stratified our analysis by physical activity status and compared the results within each stratum (physically active or physically inactive) to assess the pattern of the association between genetic liability and hypertension across the physical activity and BMI strata. To exclude any potential effect of blood pressure-lowering medication on the findings, we performed a sensitivity analysis in which we repeated our analyses excluding participants on blood pressure-lowering medication from our primary analysis (n=26,409). Within the whole sample, an interaction test was performed to determine whether the effect of genetic liability on hypertension was modulated by physical activity.

To exclude bias due to any potential effect of favourable adiposity SNPs on physical activity, we conducted a single SNP analysis to assess the effect of each favourable adiposity SNP on physical activity. We assessed this effect across the entire population, as well as within the high and low body mass subpopulations. The primary objective was to investigate whether the protective effect of favourable adiposity genetic factors against hypertension risk is influenced indirectly through physical activity. If we identified SNPs significantly associated with physical activity, we performed a sensitivity analysis in which we excluded these SNPs to create a new favourable adiposity genetic liability. We then re-examined the association between favourable adiposity genetic liability and hypertension. To understand the combined effect of genetic liability and physical activity status on the prevalence of hypertension, we selected a reference group from participants with low genetic liability and physical inactivity. Risk of hypertension in all other combinations of genetic liability and physical activity was compared with this reference group.

To additionally quantify the pure effect of physical activity on the observed effects, we compared the modulatory effect of physical activity on the association of genetic liability and hypertension within subgroups with similar BMI and genetic liability.

Statistical significance was accepted if the type 1 error (*P*-value) was below 0.05. All statistical analyses were conducted in R v4.0.0 ^36^.

## Results

This study included 230,027 UKB participants of European ancestry (Table 1). Within the whole sample studied, 144,381 had high body mass (BMI ≥ 25 kg/m^2^), while 85,646 had low body mass. Physically inactive participants had a higher BMI, with a mean of 28.2 kg/m² (SD= 5.33), compared to physically active participants, (mean BMI= 26.7 kg/m²; SD= 4.39). Among physically active participants 45.7% were hypertensive which was less than physically inactive participants (47.3%; P< 0.001).

**Table 1.**
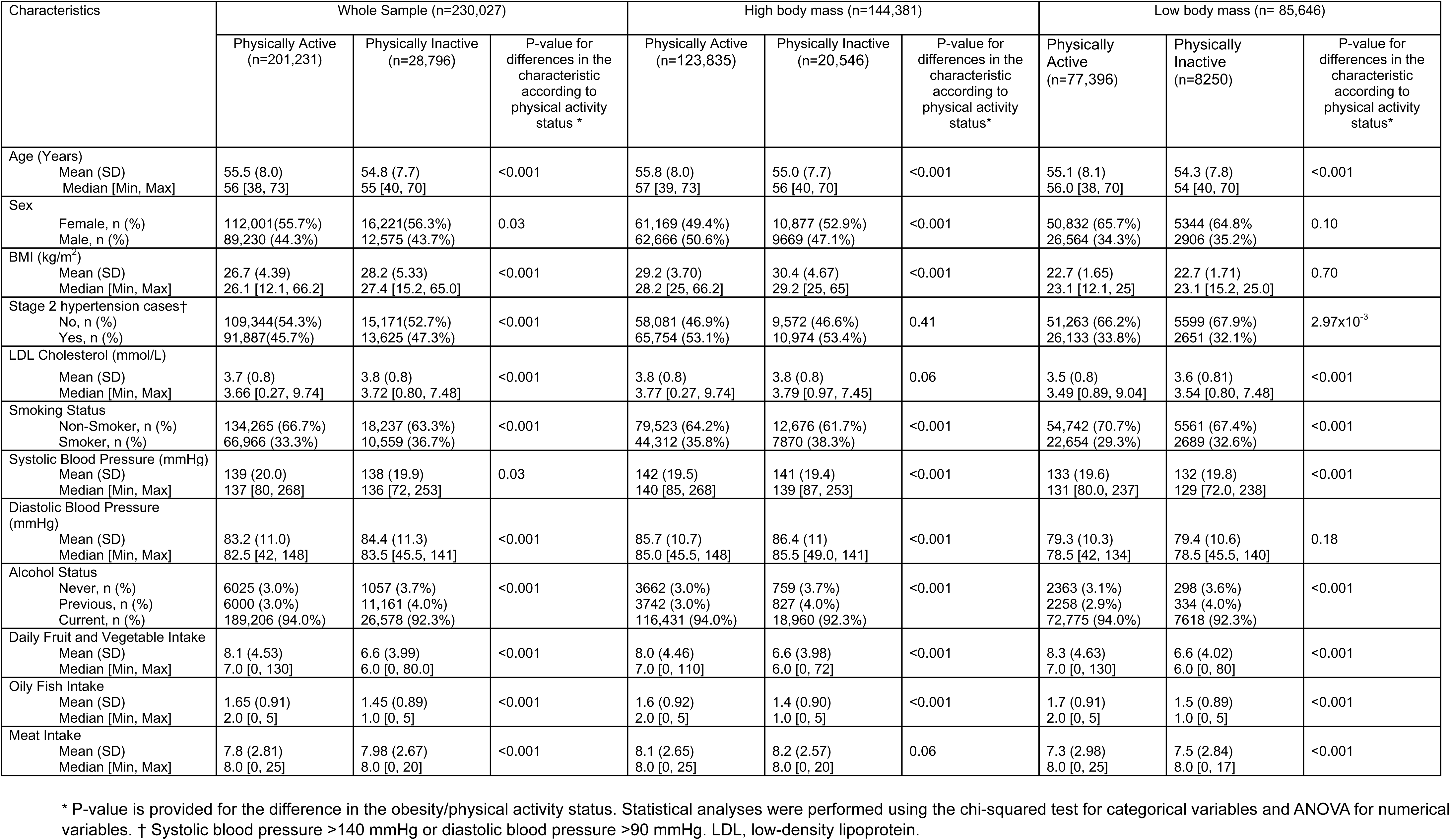
Baseline characteristics of the study sample.

| Characteristics | Whole Sample (n=230,027) |  |  | High body mass (n=144,381) |  |  | Low body mass (n= 85,646) |  |  |
| --- | --- | --- | --- | --- | --- | --- | --- | --- | --- |
|  | Physically Active<br>(n=201,231) | Physically Inactive<br>(n=28,796) | P-value for<br>differences in the<br>characteristic<br>according to<br>physical activity<br>status * | Physically Active<br>(n=123,835) | Physically Inactive<br>(n=20,546) | P-value for<br>differences in the<br>characteristic<br>according to<br>physical activity<br>status* | Physically<br>Active<br>(n=77,396) | Physically<br>Inactive<br>(n=8250) | P-value for<br>differences in the<br>characteristic<br>according to<br>physical activity<br>status* |
| Age (Years) |  |  |  |  |  |  |  |  |  |
| Mean (SD) | 55.5 (8.0) | 54.8 (7.7) | <0.001 | 55.8 (8.0) | 55.0 (7.7) | <0.001 | 55.1 (8.1) | 54.3 (7.8) | <0.001 |
| Median [Min, Max] | 56 [38, 73] | 55 [40, 70] |  | 57 [39, 73] | 56 [40, 70] |  | 56.0 [38, 70] | 54 [40, 70] |  |
| Sex |  |  |  |  |  |  |  |  |  |
| Female, n (%) | 112,001(55.7%) | 16,221(56.3%) | 0.03 | 61,169 (49.4%) | 10,877 (52.9%) | <0.001 | 50,832 (65.7%) | 5344 (64.8%) | 0.10 |
| Male, n (%) | 89,230 (44.3%) | 12,575 (43.7%) |  | 62,666 (50.6%) | 9669 (47.1%) |  | 26,564 (34.3%) | 2906 (35.2%) |  |
| BMI (kg/m<sup>2</sup>) |  |  |  |  |  |  |  |  |  |
| Mean (SD) | 26.7 (4.39) | 28.2 (5.33) | <0.001 | 29.2 (3.70) | 30.4 (4.67) | <0.001 | 22.7 (1.65) | 22.7 (1.71) | 0.70 |
| Median [Min, Max] | 26.1 [12.1, 66.2] | 27.4 [15.2, 65.0] |  | 28.2 [25, 66.2] | 29.2 [25, 65] |  | 23.1 [12.1, 25] | 23.1 [15.2, 25.0] |  |
| Stage 2 hypertension cases† |  |  |  |  |  |  |  |  |  |
| No, n (%) | 109,344(54.3%) | 15,171(52.7%) | <0.001 | 58,081 (46.9%) | 9,572 (46.6%) | 0.41 | 51,263 (66.2%) | 5599 (67.9%) | 2.97x10<sup>-3</sup> |
| Yes, n (%) | 91,887(45.7%) | 13,625 (47.3%) |  | 65,754 (53.1%) | 10,974 (53.4%) |  | 26,133 (33.8%) | 2651 (32.1%) |  |
| LDL Cholesterol (mmol/L) |  |  |  |  |  |  |  |  |  |
| Mean (SD) | 3.7 (0.8) | 3.8 (0.8) | <0.001 | 3.8 (0.8) | 3.8 (0.8) | 0.06 | 3.5 (0.8) | 3.6 (0.81) | <0.001 |
| Median [Min, Max] | 3.66 [0.27, 9.74] | 3.72 [0.80, 7.48] |  | 3.77 [0.27, 9.74] | 3.79 [0.97, 7.45] |  | 3.49 [0.89, 9.04] | 3.54 [0.80, 7.48] |  |
| Smoking Status |  |  |  |  |  |  |  |  |  |
| Non-Smoker, n (%) | 134,265 (66.7%) | 18,237 (63.3%) | <0.001 | 79,523 (64.2%) | 12,676 (61.7%) | <0.001 | 54,742 (70.7%) | 5561 (67.4%) | <0.001 |
| Smoker, n (%) | 66,966 (33.3%) | 10,559 (36.7%) |  | 44,312 (35.8%) | 7870 (38.3%) |  | 22,654 (29.3%) | 2689 (32.6%) |  |
| Systolic Blood Pressure (mmHg) |  |  |  |  |  |  |  |  |  |
| Mean (SD) | 139 (20.0) | 138 (19.9) | 0.03 | 142 (19.5) | 141 (19.4) | <0.001 | 133 (19.6) | 132 (19.8) | <0.001 |
| Median [Min, Max] | 137 [80, 268] | 136 [72, 253] |  | 140 [85, 268] | 139 [87, 253] |  | 131 [80.0, 237] | 129 [72.0, 238] |  |
| Diastolic Blood Pressure (mmHg) |  |  |  |  |  |  |  |  |  |
| Mean (SD) | 83.2 (11.0) | 84.4 (11.3) | <0.001 | 85.7 (10.7) | 86.4 (11) | <0.001 | 79.3 (10.3) | 79.4 (10.6) | 0.18 |
| Median [Min, Max] | 82.5 [42, 148] | 83.5 [45.5, 141] |  | 85.0 [45.5, 148] | 85.5 [49.0, 141] |  | 78.5 [42, 134] | 78.5 [45.5, 140] |  |
| Alcohol Status |  |  |  |  |  |  |  |  |  |
| Never, n (%) | 6025 (3.0%) | 1057 (3.7%) | <0.001 | 3662 (3.0%) | 759 (3.7%) | <0.001 | 2363 (3.1%) | 298 (3.6%) | <0.001 |
| Previous, n (%) | 6000 (3.0%) | 11,161 (4.0%) |  | 3742 (3.0%) | 827 (4.0%) |  | 2258 (2.9%) | 334 (4.0%) |  |
| Current, n (%) | 189,206 (94.0%) | 26,578 (92.3%) |  | 116,431 (94.0%) | 18,960 (92.3%) |  | 72,775 (94.0%) | 7618 (92.3%) |  |
| Daily Fruit and Vegetable Intake |  |  |  |  |  |  |  |  |  |
| Mean (SD) | 8.1 (4.53) | 6.6 (3.99) | <0.001 | 8.0 (4.46) | 6.6 (3.98) | <0.001 | 8.3 (4.63) | 6.6 (4.02) | <0.001 |
| Median [Min, Max] | 7.0 [0, 130] | 6.0 [0, 80.0] |  | 7.0 [0, 110] | 6.0 [0, 72] |  | 7.0 [0, 130] | 6.0 [0, 80] |  |
| Oily Fish Intake |  |  |  |  |  |  |  |  |  |
| Mean (SD) | 1.65 (0.91) | 1.45 (0.89) | <0.001 | 1.6 (0.92) | 1.4 (0.90) | <0.001 | 1.7 (0.91) | 1.5 (0.89) | <0.001 |
| Median [Min, Max] | 2.0 [0, 5] | 1.0 [0, 5] |  | 2.0 [0, 5] | 1.0 [0, 5] |  | 2.0 [0, 5] | 1.0 [0, 5] |  |
| Meat Intake |  |  |  |  |  |  |  |  |  |
| Mean (SD) | 7.8 (2.81) | 7.98 (2.67) | <0.001 | 8.1 (2.65) | 8.2 (2.57) | 0.06 | 7.3 (2.98) | 7.5 (2.84) | <0.001 |
| Median [Min, Max] | 8.0 [0, 25] | 8.0 [0, 20] |  | 8.0 [0, 25] | 8.0 [0, 20] |  | 8.0 [0, 25] | 8.0 [0, 17] |  |

\* P-value is provided for the difference in the obesity/physical activity status. Statistical analyses were performed using the chi-squared test for categorical variables and ANOVA for numerical variables. † Systolic blood pressure >140 mmHg or diastolic blood pressure >90 mmHg. LDL, low-density lipoprotein.

Within the high body mass group, 86% (n=123,835) participants were physically active. We observed no significant difference in the prevalence of stage 2 hypertension between physically active *vs.* physically inactive high-body mass participants (*P*=0.41).

Among low-body-mass participants, 90% (n=77,396) were physically active. Within this group, we observed no difference in the BMI between physically active and physically inactive participants (*P*=0.70). Among low body mass physically active participants, 33.8% were hypertensive which was more than physically inactive participants (32.1%; *P*=2.97x10^-3^).

### The effect of genetic liability on hypertension stratified for body mass

Among high body mass participants, the risk of hypertension was significantly reduced depending on levels of genetic liability (Adjusted OR _moderate vs. low genetic liability_= 0.95; 95% CI= 0.93,0.98; *P*=5.90 x10^-4^; Adjusted OR _high vs. low genetic liability_= 0.92; 95% CI= 0.89,0.94; p=1.26 x10^-10^) (**Table 2**). Similar results were observed within the low body mass sample (Adjusted OR _moderate vs. low genetic liability_= 0.96; 95% CI= 0.93,1.00; *P*=0.03; Adjusted OR _high vs. low genetic liability_= 0.94; 95% CI= 0.91,0.98; *P*=9.95 x10^-4^) (**Table 2**).

**Table 2:** The effect of genetic liability on hypertension stratified for body mass.

| Obesity Status | FA Genetic Liability Categories | Hypertensive (n) | Non-Hypertensive (n) | % Hypertensive | Unadjusted |  |  | Minimally Adjusted <sup>a</sup> |  |  | Adjusted <sup>b</sup> |  |  |
| --- | --- | --- | --- | --- | --- | --- | --- | --- | --- | --- | --- | --- | --- |
|  |  |  |  |  | Odds Ratio <sub>c</sub> | 95% CI <sup>d</sup> | *P*-value for Odds Ratio* | Odds Ratio <sub>c</sub> | 95% CI <sup>d</sup> | *P*-value for Odds Ratio* | Odds Ratio <sub>c</sub> | 95% CI <sup>d</sup> | *P*-value for Odds Ratio* |
| High body mass | Low | 25,552 | 21,578 | 54.22 | 1 (reference) |  |  | 1 (reference) |  |  | 1 (reference) |  |  |
|  | Moderate | 25,467 | 22,549 | 53.04 | 0.95 | 0.93 - 0.989 | 2.71 x10<sup>-4</sup> | 0.95 | 0.93 - 0.98 | 1.32 x10<sup>-4</sup> | 0.95 | 0.93 - 0.98 | 5.90 x10<sup>-4</sup> |
|  | High | 25,709 | 23,526 | 52.22 | 0.92 | 0.90 - 0.95 | 5.08 x10<sup>-10</sup> | 0.91 | 0.89 - 0.93 | 1.18 x10<sup>-12</sup> | 0.92 | 0.89 - 0.94 | 1.26 x10<sup>-10</sup> |
| Low body mass | Low | 10,165 | 19,430 | 34.35 | 1 (reference) |  |  | 1 (reference) |  |  | 1 (reference) |  |  |
|  | Moderate | 9564 | 19,071 | 33.40 | 0.96 | 0.93 - 0.99 | 1.57 x10<sup>-2</sup> | 0.96 | 0.92 - 0.99 | 1.20 x10<sup>-2</sup> | 0.96 | 0.93 - 1.00 | 0.03 |
|  | High | 9055 | 18,361 | 33.03 | 0.94 | 0.91 - 0.98 | 8.74 x10<sup>-4</sup> | 0.93 | 0.90 - 0.97 | 1.39 x10<sup>-4</sup> | 0.94 | 0.91 - 0.98 | 9.95 x10<sup>-4</sup> |

Odds ratios show the risk of prevalent stage 2 hypertension for participants belonging to each combination compared with the reference group (Low genetic liability).
Abbreviations: FA, Favourable Adiposity
a Adjusted for age and sex. b Adjusted for age, sex, smoking status, alcohol status, meat and fish intake, fruit and vegetable intake, and low-density lipoprotein cholesterol.
c , d, \*, Odds ratio, 95% confidence interval and p-value for odds ratio, are provided for the joint association between favourable adiposity genetic liability and physical activity status on the risk of hypertension. The values are derived from logistic regression models.

### The effect of genetic liability on hypertension stratified for body mass and physical activity status

**Table 3** shows the modulatory effect of physical activity on the association between favourable adiposity genetic liability and hypertension stratified by BMI. Within high body mass sample (**Table 3**), the favourable adiposity genetic liability reduced the risk of hypertension in all subgroups including within the whole sample (Adjusted OR= 0.97; 95% CI= 0.95, 0.98; *P*=1.01x10^-10^), within the physically active subgroup (Adjusted OR= 0.97; 95% CI= 0.95, 0.98; *P*=3.45x10^-9^), as well as within the physically inactive subgroups (OR= 0.96; 95% CI= 0.93, 0.99; *P*=7.80 x10^-3^). Among low body mass sample, similar results were observed for the whole sample (Adjusted OR= 0.97; 95% CI= 0.96, 0.99; *P*=1.79x10^-4^) and the physically active group (Adjusted OR= 0.97; 95% CI= 0.96, 0.99; *P*=3.60x10^-4^), but not for the physically inactive subgroup.

**Table 3.**
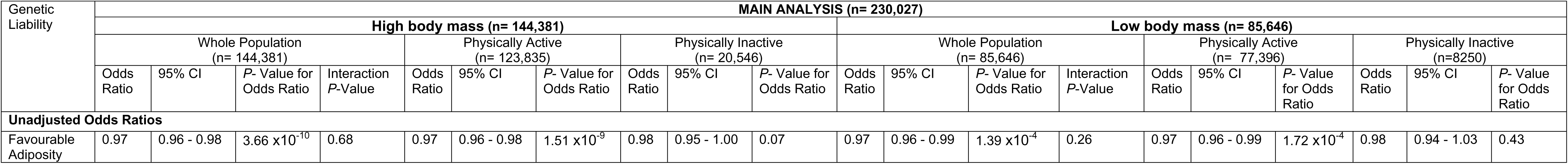
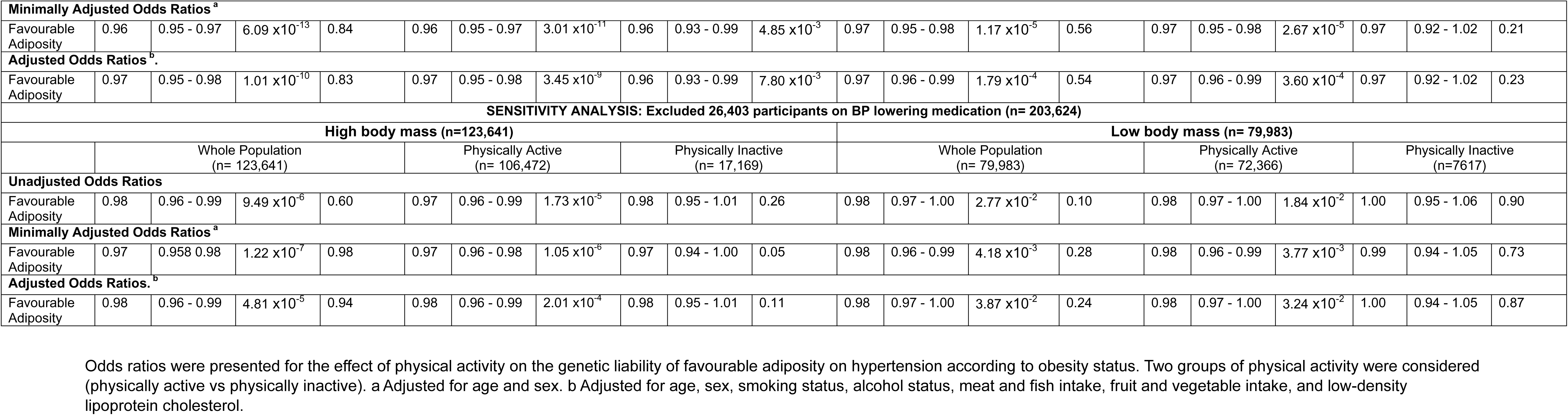
The effect of genetic liability on hypertension stratified for body mass and physical activity status.

| Genetic Liability | MAIN ANALYSIS (n= 230,027) |  |  |  |  |  |  |  |  |  |  |  |  |  |  |  |  |  |  |  |
| --- | --- | --- | --- | --- | --- | --- | --- | --- | --- | --- | --- | --- | --- | --- | --- | --- | --- | --- | --- | --- |
|  | High body mass (n= 144,381) |  |  |  |  |  |  |  |  |  | Low body mass (n= 85,646) |  |  |  |  |  |  |  |  |  |
|  | Whole Population (n= 144,381) |  |  |  | Physically Active (n= 123,835) |  |  | Physically Inactive (n= 20,546) |  |  | Whole Population (n= 85,646) |  |  |  | Physically Active (n= 77,396) |  |  | Physically Inactive (n=8250) |  |  |
|  | Odds Ratio | 95% CI | *P*- Value for Odds Ratio | Interaction *P*-Value | Odds Ratio | 95% CI | *P*- Value for Odds Ratio | Odds Ratio | 95% CI | *P*- Value for Odds Ratio | Odds Ratio | 95% CI | *P*- Value for Odds Ratio | Interaction *P*-Value | Odds Ratio | 95% CI | *P*- Value for Odds Ratio | Odds Ratio | 95% CI | *P*- Value for Odds Ratio |
| Unadjusted Odds Ratios |  |  |  |  |  |  |  |  |  |  |  |  |  |  |  |  |  |  |  |  |
| Favourable Adiposity | 0.97 | 0.96 - 0.98 | 3.66 x10<sup>-10</sup> | 0.68 | 0.97 | 0.96 - 0.98 | 1.51 x10<sup>-9</sup> | 0.98 | 0.95 - 1.00 | 0.07 | 0.97 | 0.96 - 0.99 | 1.39 x10<sup>-4</sup> | 0.26 | 0.97 | 0.96 - 0.99 | 1.72 x10<sup>-4</sup> | 0.98 | 0.94 - 1.03 | 0.43 |

|  |  |  |  |  |  |  |  |  |  |  |  |  |  |  |  |  |  |  |  |  |
| --- | --- | --- | --- | --- | --- | --- | --- | --- | --- | --- | --- | --- | --- | --- | --- | --- | --- | --- | --- | --- |
| Minimally Adjusted Odds Ratios<sup>a</sup> |  |  |  |  |  |  |  |  |  |  |  |  |  |  |  |  |  |  |  |  |
| Favourable Adiposity | 0.96 | 0.95 - 0.97 | 6.09 x10<sup>-13</sup> | 0.84 | 0.96 | 0.95 - 0.97 | 3.01 x10<sup>-11</sup> | 0.96 | 0.93 - 0.99 | 4.85 x10<sup>-3</sup> | 0.97 | 0.95 - 0.98 | 1.17 x10<sup>-5</sup> | 0.56 | 0.97 | 0.95 - 0.98 | 2.67 x10<sup>-5</sup> | 0.97 | 0.92 - 1.02 | 0.21 |
| Adjusted Odds Ratios<sup>b</sup> |  |  |  |  |  |  |  |  |  |  |  |  |  |  |  |  |  |  |  |  |
| Favourable Adiposity | 0.97 | 0.95 - 0.98 | 1.01 x10<sup>-10</sup> | 0.83 | 0.97 | 0.95 - 0.98 | 3.45 x10<sup>-9</sup> | 0.96 | 0.93 - 0.99 | 7.80 x10<sup>-3</sup> | 0.97 | 0.96 - 0.99 | 1.79 x10<sup>-4</sup> | 0.54 | 0.97 | 0.96 - 0.99 | 3.60 x10<sup>-4</sup> | 0.97 | 0.92 - 1.02 | 0.23 |
| SENSITIVITY ANALYSIS: Excluded 26,403 participants on BP lowering medication (n= 203,624) |  |  |  |  |  |  |  |  |  |  |  |  |  |  |  |  |  |  |  |  |
| High body mass (n=123,641) |  |  |  |  |  |  |  |  |  |  | Low body mass (n= 79,983) |  |  |  |  |  |  |  |  |  |
|  | Whole Population<br>(n= 123,641) |  |  |  | Physically Active<br>(n= 106,472) |  |  | Physically Inactive<br>(n= 17,169) |  |  | Whole Population<br>(n= 79,983) |  |  |  | Physically Active<br>(n= 72,366) |  |  | Physically Inactive<br>(n=7617) |  |  |
| Unadjusted Odds Ratios |  |  |  |  |  |  |  |  |  |  |  |  |  |  |  |  |  |  |  |  |
| Favourable Adiposity | 0.98 | 0.96 - 0.99 | 9.49 x10<sup>-6</sup> | 0.60 | 0.97 | 0.96 - 0.99 | 1.73 x10<sup>-5</sup> | 0.98 | 0.95 - 1.01 | 0.26 | 0.98 | 0.97 - 1.00 | 2.77 x10<sup>-2</sup> | 0.10 | 0.98 | 0.97 - 1.00 | 1.84 x10<sup>-2</sup> | 1.00 | 0.95 - 1.06 | 0.90 |
| Minimally Adjusted Odds Ratios<sup>a</sup> |  |  |  |  |  |  |  |  |  |  |  |  |  |  |  |  |  |  |  |  |
| Favourable Adiposity | 0.97 | 0.958 0.98 | 1.22 x10<sup>-7</sup> | 0.98 | 0.97 | 0.96 - 0.98 | 1.05 x10<sup>-6</sup> | 0.97 | 0.94 - 1.00 | 0.05 | 0.98 | 0.96 - 0.99 | 4.18 x10<sup>-3</sup> | 0.28 | 0.98 | 0.96 - 0.99 | 3.77 x10<sup>-3</sup> | 0.99 | 0.94 - 1.05 | 0.73 |
| Adjusted Odds Ratios.<sup>b</sup> |  |  |  |  |  |  |  |  |  |  |  |  |  |  |  |  |  |  |  |  |
| Favourable Adiposity | 0.98 | 0.96 - 0.99 | 4.81 x10<sup>-5</sup> | 0.94 | 0.98 | 0.96 - 0.99 | 2.01 x10<sup>-4</sup> | 0.98 | 0.95 - 1.01 | 0.11 | 0.98 | 0.97 - 1.00 | 3.87 x10<sup>-2</sup> | 0.24 | 0.98 | 0.97 - 1.00 | 3.24 x10<sup>-2</sup> | 1.00 | 0.94 - 1.05 | 0.87 |

Odds ratios were presented for the effect of physical activity on the genetic liability of favourable adiposity on hypertension according to obesity status. Two groups of physical activity were considered (physically active vs physically inactive). a Adjusted for age and sex. b Adjusted for age, sex, smoking status, alcohol status, meat and fish intake, fruit and vegetable intake, and low-density lipoprotein cholesterol.

As a sensitivity analysis, participants taking blood pressure lowering medication were excluded. Among individuals with high body mass, the results stayed similar for the whole sample and the physically active group, but not for the physically inactive subgroup (*P*=0.11; **Table 3).** The results were slightly attenuated among individuals with low body mass both within the whole sample (Adjusted OR= 0.98; 95% CI= 0.97, 1.00; *P*=3.87 x10^-2^) and within the physically active sample (OR= 0.98; 95% CI= 0.97, 1.00; *P*=3.24 x10^-2^; **Table 3).**

### The effect of favourable adiposity SNPs on physical activity

We investigated whether the protective effect of favourable adiposity SNPs on BMI and therefore hypertension could potentially be driven by physical activity. To explore this, we examined the impact of all favourable adiposity-related SNPs on physical activity (**Table 4**) and BMI (**Supplementary Table 3**). Of the 35 favourable adiposity SNPs used in estimating the favourable adiposity genetic liability, rs987469 was associated with reduced physical activity within the whole sample (Adjusted β = - 12.04 per copy C allele; 95% CI= -23.22, -0.87; *P*=0.03) and within the high body mass sample (Adjusted β = -18.01 per copy C allele; 95% CI= -32.10, -3.91; *P*=0.01). Additionally, rs12369179 (Adjusted β = -14.90 per copy C allele; 95% CI= - 28.81, -0.99; *P*=0.04) and rs573454216 (Adjusted β = -15.99 per copy G allele; 95% CI= -30.11, -1.88; *P*=0.03) were associated with reduced physical activity within the high body mass sample. A statistically significant association was also observed in the relationship between rs4684847 and increased physical activity within the whole sample (Adjusted β = 15.89 per copy C allele; 95% CI= 4.85, 26.93; *P*=4.79x10^-3^) and within the high body mass sample (Adjusted β = 18.77 per copy C allele; 95% CI= 4.88, 32.65; *P*=8.06x10^-3^) (**Table 4**). For rs12369179, rs4684847, and rs987469 the allele that reduced physical activity increased BMI (**Supplementary Table 3**).

**Table 4.** Effect of favourable adiposity SNPs and genetic liability on the total amount of MET/minute physical activity.

| Favourable Adiposity SNPs<br>(Coded allele) | Whole Population<br>(N= 230,027) |  |  | High body mass Population<br>(N= 144,381) |  |  | Low body mass Population<br>(N= 85,646) |  |  |
| --- | --- | --- | --- | --- | --- | --- | --- | --- | --- |
|  | Effect estimate (β) | 95% CI | P- value | Effect estimate (β) | 95% CI | P- value | Effect estimate (β) | 95% CI | P- value |
| rs12369179<sup>Ω</sup> (C) |  |  |  |  |  |  |  |  |  |
| Unadjusted Odds Ratios | -10.13 | -21.11 – 0.84 | 0.07 | -14.26 | -28.23 – -0.30 | 0.04 | -0.17 | -17.85 – 17.52 | 0.99 |
| Adjusted Odds Ratios<sup>†</sup> | -10.63 | -21.57 – 0.32 | 0.06 | -14.90 | -28.81 – -0.99 | 0.04 | 0.13 | -17.52 – 17.78 | 0.99 |
| rs4684847<sup>§</sup> (C) |  |  |  |  |  |  |  |  |  |
| Unadjusted Odds Ratios | 16.36 | 5.29 – 27.43 | 3.77x10<sup>-3</sup> | 18.92 | 4.98 – 32.86 | 7.81x10<sup>-3</sup> | 13.24 | -4.91 – 31.39 | 0.15 |
| Adjusted Odds Ratios<sup>†</sup> | 15.89 | 4.85 – 26.93 | 4.79x10<sup>-3</sup> | 18.77 | 4.88 – 32.65 | 8.06x10<sup>-3</sup> | 12.66 | -5.46 – 30.77 | 0.17 |
| rs62271373 (T) |  |  |  |  |  |  |  |  |  |
| Unadjusted Odds Ratios | -5.15 | -16.17 – 5.88 | 0.36 | -8.31 | -22.21 – 5.59 | 0.24 | 0.50 | -17.56 – 18.57 | 0.96 |
| Adjusted Odds Ratios<sup>†</sup> | -5.85 | -16.85 – 5.14 | 0.30 | -8.81 | -22.65 – 5.03 | 0.21 | -0.35 | -18.37 – 17.67 | 0.97 |
| rs72959041 (G) |  |  |  |  |  |  |  |  |  |
| Unadjusted Odds Ratios | -1.75 | -12.71 – 9.22 | 0.76 | -1.11 | -15.00 – 12.79 | 0.88 | -1.42 | -19.23 – 16.40 | 0.88 |
| Adjusted Odds Ratios<sup>†</sup> | -1.34 | -12.28 – 9.60 | 0.81 | -0.57 | -14.40 – 13.27 | 0.94 | -0.86 | -18.64 – 16.91 | 0.92 |
| rs12130231 (A) |  |  |  |  |  |  |  |  |  |
| Unadjusted Odds Ratios | -9.69 | -20.95 – 1.57 | 0.09 | -13.44 | -27.65 – 0.77 | 0.06 | -1.49 | -19.92 – 16.93 | 0.87 |
| Adjusted Odds Ratios<sup>†</sup> | -10.17 | -21.40 – 1.06 | 0.08 | -13.61 | -27.76 – 0.54 | 0.06 | -2.29 | -20.67 – 16.10 | 0.81 |
| rs7133378 (G) |  |  |  |  |  |  |  |  |  |
| Unadjusted Odds Ratios | 5.60 | -5.65 – 16.84 | 0.33 | -0.71 | -14.86 – 13.45 | 0.92 | 17.63 | -0.84 – 36.09 | 0.06 |
| Adjusted Odds Ratios<sup>†</sup> | 5.19 | -6.03 – 16.41 | 0.37 | -0.14 | -14.24 – 13.96 | 0.98 | 16.44 | -1.99 – 34.86 | 0.08 |
| rs4821764 (G) |  |  |  |  |  |  |  |  |  |
| Unadjusted Odds Ratios | -2.52 | -13.72 – 8.69 | 0.66 | -7.59 | -21.71 – 6.52 | 0.29 | 7.68 | -10.69 – 26.04 | 0.41 |
| Adjusted Odds Ratios<sup>†</sup> | -2.31 | -13.49 – 8.86 | 0.69 | -7.17 | -21.23 – 6.88 | 0.32 | 8.26 | -10.07 – 26.58 | 0.38 |
| rs13389219 (C) |  |  |  |  |  |  |  |  |  |
| Unadjusted Odds Ratios | 3.83 | -7.44 – 15.09 | 0.51 | 1.90 | -12.31 – 16.12 | 0.79 | 7.36 | -11.08 – 25.79 | 0.43 |
| Adjusted Odds Ratios<sup>†</sup> | 3.50 | -7.74 – 14.73 | 0.54 | 2.52 | -11.64 – 16.67 | 0.73 | 6.25 | -12.15 – 24.65 | 0.51 |
| rs2943653 (C) |  |  |  |  |  |  |  |  |  |
| Unadjusted Odds Ratios | 2.61 | -8.60 – 13.81 | 0.65 | 4.09 | -10.06 – 18.24 | 0.57 | -0.07 | -18.38 – 18.23 | 0.99 |
| Adjusted Odds Ratios<sup>†</sup> | 1.88 | -9.29 – 13.06 | 0.74 | 2.90 | -11.19 – 16.99 | 0.69 | -0.18 | -18.44 – 18.08 | 0.99 |
| rs7258937 (C) |  |  |  |  |  |  |  |  |  |
| Unadjusted Odds Ratios | -0.82 | -12.02 – 10.39 | 0.89 | -0.85 | -15.00 – 13.29 | 0.91 | 0.81 | -17.50 – 19.13 | 0.93 |
| Adjusted Odds Ratios<sup>†</sup> | -1.45 | -12.62 – 9.73 | 0.80 | -1.17 | -15.26 – 12.91 | 0.87 | 0.04 | -18.24 – 18.32 | 1.00 |
| rs72697297 (T) |  |  |  |  |  |  |  |  |  |
| Unadjusted Odds Ratios | -1.40 | -12.47 – 9.67 | 0.80 | -3.80 | -17.76 – 10.16 | 0.59 | 2.46 | -15.68 – 20.59 | 0.79 |
| Adjusted Odds Ratios<sup>†</sup> | -1.43 | -12.47 – 9.62 | 0.80 | -2.73 | -16.63 – 11.17 | 0.70 | 0.99 | -17.11 – 19.09 | 0.92 |
| rs972283 (A) |  |  |  |  |  |  |  |  |  |
| Unadjusted Odds Ratios | -0.76 | -11.99 – 10.48 | 0.90 | -0.03 | -14.20 – 14.14 | 1.00 | 0.85 | -17.56 – 19.25 | 0.93 |
| Adjusted Odds Ratios<sup>†</sup> | -1.25 | -12.46 – 9.96 | 0.83 | -0.60 | -14.71 – 13.514 | 0.93 | 1.11 | -17.25 – 19.48 | 0.91 |
| rs11135038 (T) |  |  |  |  |  |  |  |  |  |
| Unadjusted Odds Ratios | -7.41 | -18.55 – 3.73 | 0.19 | -9.74 | -23.79 – 4.30 | 0.17 | -2.23 | -20.48 – 16.02 | 0.81 |
| Adjusted Odds Ratios<sup>†</sup> | -7.97 | -19.08 – 3.14 | 0.16 | -9.74 | -23.73 – 4.25 | 0.17 | -2.92 | -21.13 – 15.29 | 0.75 |
| rs9851766 (A) |  |  |  |  |  |  |  |  |  |
| Unadjusted Odds Ratios | -5.18 | -16.27 – 5.91 | 0.36 | -5.48 | -19.53 – 8.57 | 0.45 | -1.55 | -19.57 – 16.47 | 0.87 |
| Adjusted Odds Ratios<sup>†</sup> | -5.45 | -16.51 – 5.61 | 0.33 | -5.39 | -19.38 – 8.60 | 0.45 | -1.55 | -19.53 – 16.43 | 0.87 |
| rs142186653 (A) |  |  |  |  |  |  |  |  |  |
| Unadjusted Odds Ratios | -1.68 | -12.98 – 9.62 | 0.77 | -0.20 | -14.43 – 14.02 | 0.98 | -2.73 | -21.28 – 15.81 | 0.77 |
| Adjusted Odds Ratios<sup>†</sup> | -2.48 | -13.75 – 8.79 | 0.67 | -0.89 | -15.05 – 13.28 | 0.90 | -3.35 | -21.86 – 15.15 | 0.72 |

Table 4. Effect of favourable adiposity SNPs and genetic liability on physical activity (continued)
| Favourable Adiposity SNPS | Whole Population (N= 230,027) |  |  | High body mass Population (N= 144,381) |  |  | Low body mass Population (N= 85,646) |  |  |
| --- | --- | --- | --- | --- | --- | --- | --- | --- | --- |
|  | Effect estimate (β) | 95% CI | P- value | Effect estimate (β) | 95% CI | P- value | Effect estimate (β) | 95% CI | P- value |
| rs987469<sup>Ω</sup> (C) |  |  |  |  |  |  |  |  |  |
| Unadjusted Odds Ratios | -12.01 | -23.22 – -0.81 | 0.04 | -17.45 | -31.60 – -3.29 | 0.02 | -2.16 | -20.44 – 16.12 | 0.82 |
| Adjusted Odds Ratios<sup>†</sup> | -12.04 | -23.22 – -0.87 | 0.03 | -18.01 | -32.10 – -3.91 | 0.01 | -1.27 | -19.51 – 16.98 | 0.89 |
| rs2980888 (T) |  |  |  |  |  |  |  |  |  |
| Unadjusted Odds Ratios | -5.50 | -16.70 – 5.71 | 0.34 | -6.04 | -20.21 – 8.13 | 0.40 | -0.38 | -18.62 – 17.86 | 0.97 |
| Adjusted Odds Ratios<sup>†</sup> | -6.62 | -17.79 – 4.55 | 0.25 | -6.73 | -20.84 – 7.39 | 0.35 | -1.19 | -19.39 – 17.01 | 0.90 |
| rs30351 (G) |  |  |  |  |  |  |  |  |  |
| Unadjusted Odds Ratios | 7.43 | -4.14 – 19.00 | 0.21 | 8.43 | -6.16 – 23.01 | 0.26 | 5.98 | -12.96 – 24.93 | 0.54 |
| Adjusted Odds Ratios<sup>†</sup> | 6.50 | -5.04 –18.04 | 0.27 | 7.09 | -7.44 – 21.61 | 0.34 | 4.95 | -13.96 – 23.85 | 0.61 |
| rs12681990 (T) |  |  |  |  |  |  |  |  |  |
| Unadjusted Odds Ratios | -4.36 | -15.89 – 7.17 | 0.46 | -13.68 | -28.24 – 0.89 | 0.07 | 12.01 | -6.82 – 30.84 | 0.21 |
| Adjusted Odds Ratios<sup>†</sup> | -4.88 | -16.38 – 6.62 | 0.41 | -13.80 | -28.31 – 0.70 | 0.06 | 11.17 | -7.62 – 29.96 | 0.24 |
| rs6977416 (G) |  |  |  |  |  |  |  |  |  |
| Unadjusted Odds Ratios | 2.65 | -8.69 – 14.00 | 0.65 | -0.74 | -15.05 – 13.57 | 0.92 | 8.36 | -10.20 – 26.92 | 0.38 |
| Adjusted Odds Ratios<sup>†</sup> | 2.63 | -8.69 – 13.94 | 0.65 | -1.10 | -15.35 – 13.15 | 0.88 | 8.49 | -10.03 – 27.01 | 0.37 |
| rs4976033 (A) |  |  |  |  |  |  |  |  |  |
| Unadjusted Odds Ratios | -6.59 | -17.80 – 4.62 | 0.25 | -13.46 | -27.59 – 0.66 | 0.06 | 6.34 | -12.04 – 24.72 | 0.50 |
| Adjusted Odds Ratios<sup>†</sup> | -6.69 | -17.87 – 4.49 | 0.24 | -13.77 | -27.84 – 0.29 | 0.06 | 6.85 | -11.49 – 25.19 | 0.46 |
| rs12441543 (G) |  |  |  |  |  |  |  |  |  |
| Unadjusted Odds Ratios | 0.87 | -10.39 – 12.13 | 0.88 | 2.24 | -11.95 – 16.44 | 0.76 | -0.36 | -18.82 – 18.09 | 0.97 |
| Adjusted Odds Ratios<sup>†</sup> | 0.24 | -10.99 – 11.47 | 0.97 | 1.36 | -12.77 – 15.49 | 0.85 | -0.78 | -19.19 – 17.64 | 0.93 |
| rs12940684 (C) |  |  |  |  |  |  |  |  |  |
| Unadjusted Odds Ratios | 5.08 | -6.22 – 16.39 | 0.38 | 0.65 | -13.58 – 14.89 | 0.93 | 14.50 | -4.07 – 33.07 | 0.13 |
| Adjusted Odds Ratios<sup>†</sup> | 4.48 | -6.80 – 15.76 | 0.44 | 0.03 | -14.15 – 14.21 | 1.00 | 14.10 | -4.43 – 32.63 | 0.14 |
| rs113222038 (C) |  |  |  |  |  |  |  |  |  |
| Unadjusted Odds Ratios | -8.02 | -19.15 – 3.11 | 0.16 | -7.06 | -21.11 – 6.99 | 0.33 | -9.77 | -27.94 – 8.40 | 0.29 |
| Adjusted Odds Ratios<sup>†</sup> | -7.40 | -18.49 – 3.70 | 0.19 | -6.27 | -20.27 – 7.72 | 0.38 | -9.17 | -27.30 – 8.96 | 0.32 |
| rs11045172 (A) |  |  |  |  |  |  |  |  |  |
| Unadjusted Odds Ratios | 3.66 | -7.41 – 14.74 | 0.52 | 1.34 | -12.60 – 15.28 | 0.85 | 9.09 | -9.09 – 27.27 | 0.33 |
| Adjusted Odds Ratios<sup>†</sup> | 3.68 | -7.36 – 14.73 | 0.51 | 1.80 | -12.08 – 15.69 | 0.80 | 8.85 | -9.29 – 26.99 | 0.34 |
| rs2802774 (C) |  |  |  |  |  |  |  |  |  |
| Unadjusted Odds Ratios | -1.83 | -12.97 – 9.32 | 0.75 | -1.78 | -15.83 – 12.28 | 0.80 | 0.08 | -18.17 – 18.33 | 0.99 |
| Adjusted Odds Ratios<sup>†</sup> | -2.00 | -13.11 – 9.12 | 0.72 | -2.38 | -16.38 – 11.61 | 0.74 | 0.87 | -17.33 – 19.08 | 0.93 |
| rs7233512 (G) |  |  |  |  |  |  |  |  |  |
| Unadjusted Odds Ratios | 3.74 | -7.53 – 15.00 | 0.52 | 2.39 | -11.83 – 16.61 | 0.74 | 8.31 | -10.10 – 26.72 | 0.38 |
| Adjusted Odds Ratios<sup>†</sup> | 3.34 | -7.89 – 14.58 | 0.56 | 2.47 | -11.69 – 16.63 | 0.73 | 7.78 | -10.59 – 26.15 | 0.41 |
| rs9764678 (T) |  |  |  |  |  |  |  |  |  |
| Unadjusted Odds Ratios | -0.52 | -11.72 – 10.68 | 0.92 | 6.41 | -7.69 – 20.51 | 0.37 | -12.14 | -30.54 – 6.25 | 0.20 |
| Adjusted Odds Ratios<sup>†</sup> | 0.07 | -11.11 – 11.24 | 0.99 | 6.93 | -7.12 – 20.97 | 0.33 | -11.29 | -29.65 – 7.06 | 0.23 |
| rs10876529 (T) |  |  |  |  |  |  |  |  |  |
| Unadjusted Odds Ratios | -6.98 | -18.13 – 4.17 | 0.22 | -2.83 | -16.89 – 11.23 | 0.69 | -13.60 | -31.87 – 4.68 | 0.15 |
| Adjusted Odds Ratios<sup>†</sup> | -6.70 | -17.82 – 4.43 | 0.24 | -1.99 | -15.98 – 12.01 | 0.78 | -13.22 | -31.45 – 5.01 | 0.16 |
| rs11664106 (A) |  |  |  |  |  |  |  |  |  |
| Unadjusted Odds Ratios | 0.30 | -10.84 – 11.44 | 0.96 | 2.23 | -11.81 – 16.28 | 0.76 | -1.04 | -19.29 – 17.20 | 0.91 |
| Adjusted Odds Ratios<sup>†</sup> | 0.15 | -10.96 – 11.26 | 0.98 | 2.77 | -11.22 – 16.76 | 0.70 | -1.33 | -19.53 – 16.88 | 0.89 |

Table 4. Effect of favourable adiposity SNPs and genetic liability on the total amount of MET/minute physical activity (continued)
| Favourable Adiposity SNPs | Whole Population (N= 230,027) |  |  | High body mass Population (N= 144,381) |  |  | Low body mass Population (N= 85,646) |  |  |
| --- | --- | --- | --- | --- | --- | --- | --- | --- | --- |
|  | Effect estimate (β) | 95% CI | P- value | Effect estimate (β) | 95% CI | P- value | Effect estimate (β) | 95% CI | P- value |
| rs998584 (C) |  |  |  |  |  |  |  |  |  |
| Unadjusted Odds Ratios | -2.05 | -13.30 – 9.19 | 0.72 | 0.77 | -13.41 –14.95 | 0.92 | -4.84 | -23.25 – 13.57 | 0.61 |
| Adjusted Odds Ratios<sup>†</sup> | -3.10 | -14.31 – 8.12 | 0.59 | -0.29 | -14.41 – 13.83 | 0.97 | -5.80 | -24.17 – 12.57 | 0.54 |
| rs6029180 (A) |  |  |  |  |  |  |  |  |  |
| Unadjusted Odds Ratios | 10.18 | -0.95 – 21.31 | 0.07 | 7.67 | -6.33 – 21.67 | 0.28 | 17.66 | -0.65 – 35.97 | 0.06 |
| Adjusted Odds Ratios<sup>†</sup> | 10.15 | -0.96 – 21.25 | 0.07 | 8.08 | -5.86 – 22.02 | 0.26 | 17.79 | -0.48 – 36.06 | 0.06 |
| rs13132853 (A) |  |  |  |  |  |  |  |  |  |
| Unadjusted Odds Ratios | 4.54 | -6.63 – 15.70 | 0.43 | -1.41 | -15.50 – 12.68 | 0.84 | 15.24 | -3.01 – 33.50 | 0.10 |
| Adjusted Odds Ratios<sup>†</sup> | 4.82 | -6.31 – 15.96 | 0.40 | -1.55 | -15.58 – 12.48 | 0.83 | 16.10 | -2.12 – 34.32 | 0.08 |
| rs573454216<sup>Ω</sup> (G) |  |  |  |  |  |  |  |  |  |
| Unadjusted Odds Ratios | -10.63 | -21.87 – 0.61 | 0.06 | -15.53 | -29.70 – -1.36 | 0.03 | -1.95 | -20.34 – 16.45 | 0.84 |
| Adjusted Odds Ratios<sup>†</sup> | -10.63 | -21.84 – 0.58 | 0.06 | -15.99 | -30.11 – -1.88 | 0.03 | -1.31 | -19.67 – 17.05 | 0.89 |
| rs4450871 (A) |  |  |  |  |  |  |  |  |  |
| Unadjusted Odds Ratios | -3.47 | -14.73 – 7.78 | 0.55 | -6.61 | -20.81 – 7.59 | 0.36 | 2.50 | -15.90 – 20.90 | 0.79 |
| Adjusted Odds Ratios<sup>†</sup> | -3.38 | -14.61 – 7.84 | 0.56 | -6.91 | -21.05 – 7.24 | 0.34 | 3.49 | -14.88 – 21.85 | 0.71 |
| Favourable Adiposity genetic liability |  |  |  |  |  |  |  |  |  |
| Unadjusted Odds Ratios | -6.07 | -17.39 – 5.26 | 0.29 | -15.39 | -29.67 – -1.11 | 0.03 | 17.51 | -1.04 – 36.07 | 0.06 |
| Adjusted Odds Ratios<sup>†</sup> | -7.82 | -19.12 – 3.47 | 0.18 | -16.18 | -30.40 – -1.96 | 0.03 | 16.34 | -2.17 – 34.86 | 0.08 |
| Favourable Adiposity genetic liability<sup>δ</sup> |  |  |  |  |  |  |  |  |  |
| Unadjusted Odds Ratios | -5.29 | -16.72 – 6.15 | 0.37 | -13.71 | -28.12 – 0.71 | 0.06 | 16.19 | -2.56 – 34.95 | 0.09 |
| Adjusted Odds Ratios<sup>†</sup> | -6.96 | -18.36 – 4.45 | 0.23 | -14.21 | -28.57 – 0.15 | 0.05 | 14.74 | -3.97 – 33.45 | 0.12 |

Effect estimates (β) and 95% confidence interval (CI) for favourable adiposity SNPs and favourable adiposity genetic liability are estimates from linear regression models. <sup>†</sup> Adjusted for age and sex. <sup>§</sup> Favourable adiposity SNPs associated with increased physical activity from the linear regression model. <sup>Ω</sup> Favourable adiposity SNPs associated with reduced physical activity from the linear regression model. <sup>δ</sup> Favourable adiposity genetic liability estimated without the physical activity-associated SNPs.

We observed that the genetic liability was not associated with physical activity within the whole sample or the low body mass subgroups. However, the genetic liability was associated with reduced physical activity within the high body mass sample (Adjusted β = -16.18; 95% CI= -30.40, -1.96; *P*=0.03). After excluding the physical activity-related SNPs identified in our previous step (rs12369179, rs4684847, rs987469 and rs573454216), no association between genetic liability and physical activity was observed within the whole sample or any of the subgroups (**Table 4**).

We repeated the main association analyses between genetic liability and hypertension (stratified by physical activity and BMI) excluding the four physical activity-related SNPs and observed no remarkable changes in the findings (**Supplementary Table 4**).

### Joint effects of genetic liability and physical activity on hypertension risk stratified by the BMI (excluding physical activity-related SNPs)

Stratified by BMI, the joint effect of the genetic liability and physical activity on risk of hypertension was compared with a reference group that included individuals who had a combination of low genetic liability and low physical activity as both of these factors are not optimal for risk of hypertension (**Table 5**). Among high body mass participants, all combinations of genetic liability and physical activity were associated with reduced risks of hypertension compared with the reference group (Adjusted OR _low genetic liability, physically active_ = 0.91; 95% CI= 0.86, 0.96; *P*=5.05 x10^-4^; Adjusted OR _moderate genetic liability, physically active_ = 0.87; 95% CI= 0.83, 0.92 ; *P*=8.59 x10^-7^; Adjusted OR _moderate genetic liability, physically inactive_ = 0.93; 95% CI= 0.86, 0.99; *P*=2.99 x10^-2^ ; Adjusted OR _high genetic liability, physically active_ = 0.84; 95% CI=0.80, 0.89; *P*=2.27 x10^-10^; Adjusted OR _high genetic liability, physically inactive_ = 0.91; 95% CI= 0.85, 0.97; *P*=6.80 x10^-3^; **Table 5** ; **Supplementary Table 5**). Among low body mass participants, no association was observed between any combinations of genetic liability and physical activity status with the risk of hypertension (**Table 5; Supplementary Table 5**).

**Table 5:** Joint effects of genetic liability and physical activity on hypertension risk stratified by the BMI (excluding physical activity-related SNPs^§^)

| Obesity Status | FA Genetic Liability <sup>‡</sup> Categories | Physical Activity Status | Hypertensive (n) | Non-Hypertensive (n) | % Hypertensive | Unadjusted |  |  | Minimally Adjusted <sup>a</sup> |  |  | Adjusted <sup>b</sup> |  |  |
| --- | --- | --- | --- | --- | --- | --- | --- | --- | --- | --- | --- | --- | --- | --- |
|  |  |  |  |  |  | Odds Ratio <sup>c</sup> | 95% CI <sup>d</sup> | P-value for Odds Ratio<sup>*</sup> | Odds Ratio <sup>c</sup> | 95% CI <sup>d</sup> | P-value for Odds Ratio<sup>*</sup> | Odds Ratio <sup>c</sup> | 95% CI <sup>d</sup> | P-value for Odds Ratio<sup>*</sup> |
| High body mass | Low | Physically Inactive | 3,650 | 3,055 | 54.44 | 1 (reference) |  |  | 1 (reference) |  |  | 1 (reference) |  |  |
|  | Low | Physically Active | 22,248 | 18,921 | 54.04 | 0.98 | 0.93 - 1.04 | 0.55 | 0.90 | 0.86 - 0.95 | 2.17 x10<sup>-4</sup> | 0.91 | 0.86 - 0.96 | 5.05 x10<sup>-4</sup> |
|  | Moderate | Physically Inactive | 3,589 | 3,180 | 53.02 | 0.95 | 0.88 - 1.01 | 9.94 x10<sup>-2</sup> | 0.92 | 0.86 - 0.99 | 1.86 x10<sup>-2</sup> | 0.93 | 0.86 - 0.99 | 2.99 x10<sup>-2</sup> |
|  | Moderate | Physically Active | 21,741 | 19,225 | 53.07 | 0.95 | 0.90 - 1.00 | 0.04 | 0.86 | 0.82 - 0.91 | 1.11 x10<sup>-7</sup> | 0.87 | 0.83 - 0.92 | 8.59 x10<sup>-7</sup> |
|  | High | Physically Inactive | 3,735 | 3,337 | 52.81 | 0.94 | 0.88 - 1.00 | 5.62 x10<sup>-2</sup> | 0.90 | 0.84 - 0.97 | 3.44 x10<sup>-3</sup> | 0.91 | 0.85 - 0.97 | 6.80 x10<sup>-3</sup> |
|  | High | Physically Active | 21,765 | 19,935 | 52.19 | 0.91 | 0.87 - 0.96 | 6.43 x10<sup>-4</sup> | 0.83 | 0.78 - 0.87 | 5.80 x10<sup>-12</sup> | 0.84 | 0.80 - 0.89 | 2.27 x10<sup>-10</sup> |
| Low body mass | Low | Physically Inactive | 941 | 1,996 | 32.04 | 1 (reference) |  |  | 1 (reference) |  |  | 1 (reference) |  |  |
|  | Low | Physically Active | 9,220 | 17,633 | 34.34 | 1.11 | 1.02 - 1.20 | 1.28 x10<sup>-2</sup> | 1.02 | 0.93 - 1.11 | 0.73 | 1.04 | 0.95 - 1.13 | 0.41 |
|  | Moderate | Physically Inactive | 876 | 1,776 | 33.03 | 1.05 | 0.94 - 1.17 | 0.43 | 1.01 | 0.90 - 1.13 | 0.90 | 1.00 | 0.89 - 1.13 | 0.95 |
|  | Moderate | Physically Active | 8,713 | 17,005 | 33.88 | 1.09 | 1.00 - 1.18 | 4.57 x10<sup>-2</sup> | 0.99 | 0.91 - 1.08 | 0.789 | 1.01 | 0.93 - 1.11 | 0.75 |
|  | High | Physically Inactive | 834 | 1,827 | 31.34 | 0.97 | 0.87 - 1.08 | 0.58 | 0.92 | 0.82 - 1.04 | 0.18 | 0.93 | 0.83 - 1.05 | 0.25 |
|  | High | Physically Active | 8,200 | 16,625 | 33.03 | 1.05 | 0.96 - 1.14 | 0.28 | 0.95 | 0.87 - 1.03 | 0.23 | 0.98 | 0.90 - 1.07 | 0.63 |

Odds ratios show the risk of prevalent stage 2 hypertension for participants belonging to each combination compared with the reference group (Low genetic liability combined with physically Inactive).
Abbreviations: FA, Favourable Adiposity (≠ Estimated genetic liability without rs12369179, rs4684847, rs987469 and rs573454216).
a Adjusted for age and sex. b Adjusted for age, sex, smoking status, alcohol status, meat and fish intake, fruit and vegetable intake, and low-density lipoprotein cholesterol.
c , d, \*, Odds ratio, 95% confidence interval and p-value for odds ratio, are provided for the joint association between favourable adiposity genetic liability and physical activity status on the risk of hypertension. The values are derived from logistic regression models.

We also observed that physical activity reduced risk of hypertension beyond genetic liability among high body mass individuals. In brief, the effect of the combination of low genetic liability and physical activity was significantly lower than the combination of low genetic liability and physical inactivity (OR= 0.93, 95% CI= 0.88, 0.99; *P*=1.58 x10^-2^). Similar results were observed for the combination of moderate genetic liability and physical activity vs. the combination of moderate genetic liability and physical inactivity (OR=0.91, 95% CI= 0.86, 0.96; *P*=2.91 x10^-4^) as well as the combination of high genetic liability and physical activity vs. the combination of high genetic liability and physical inactivity (OR= 0.94, 95% CI= 0.89, 0.99; *P*=1.42 x10^-2^) (**Table 6**). In contrast, within the low body mass participants no association was found in terms of physical activity reducing the risk of hypertension across any level of genetic liability (**Table 6**).

**Table 6:** Influence of Physical Activity on Hypertension Risk Across different categories of favourable Adiposity Genetic Liability.

| Obesity Status | FA Genetic Liability Categories | Physical Activity Status | genetic liability includes all SNPs Adjusted <sup>b</sup> |  |  | Sensitivity analysis excluding physical activity SNPs Adjusted <sup>b</sup> |  |  |
| --- | --- | --- | --- | --- | --- | --- | --- | --- |
|  |  |  | Odds Ratio <sup>c</sup> | 95% CI <sup>d</sup> | *P*-value <sup>*</sup> | Odds Ratio <sup>c</sup> | 95% CI <sup>d</sup> | *P*-value <sup>*</sup> |
| High body mass | Low | Physically Inactive | 1 (reference) | -- | -- | 1 (reference) | -- | -- |
| | Low | Physically Active | 0.93 | 0.88 - 0.99 | $1.58 \times 10^{-2}$ | 0.91 | 0.86 - 0.96 | $8.79 \times 10^{-4}$ |
|  | Moderate | Physically Inactive | 1 (reference) | -- | -- | 1 (reference) | -- | -- |
| | Moderate | Physically Active | 0.91 | 0.86 - 0.96 | $2.91 \times 10^{-4}$ | 0.94 | 0.89 - 0.99 | $2.89 \times 10^{-2}$ |
|  | High | Physically Inactive | 1 (reference) | -- | -- | 1 (reference) | -- | -- |
| | High | Physically Active | 0.94 | 0.89 - 0.99 | $1.42 \times 10^{-2}$ | 0.92 | 0.88 - 0.97 | $3.48 \times 10^{-3}$ |
| Low body mass | Low | Physically Inactive | 1 (reference) | -- | -- | 1 (reference) | -- | -- |
|  | Low | Physically Active | 1.06 | 0.97 - 1.15 | 0.22 | 1.04 | 0.95 - 1.14 | 0.37 |
|  | Moderate | Physically Inactive | 1 (reference) | -- | -- | 1 (reference) | -- | -- |
|  | Moderate | Physically Active | 1.04 | 0.95 - 1.14 | 0.43 | 1.01 | 0.93 - 1.11 | 0.77 |
|  | High | Physically Inactive | 1 (reference) | -- | -- | 1 (reference) | -- | -- |
|  | High | Physically Active | 1.01 | 0.92 - 1.10 | 0.90 | 1.05 | 0.95 - 1.15 | 0.33 |

Odds ratios show the risk of prevalent stage 2 hypertension for participants belonging to each combination compared with the reference group (Low genetic liability combined with physically Inactive).
Abbreviations: FA, Favourable Adiposity
<sup>b</sup> Adjusted for age, sex, smoking status, alcohol status, meat and fish intake, fruit and vegetable intake, and low-density lipoprotein cholesterol.
<sup>c</sup> , <sup>d</sup> , <sup>\*</sup> , Odds ratio, 95% confidence interval and *p*-value for odds ratio, are provided for difference in physical activity status for each category of genetic liability on the risk of hypertension. The values are derived from logistic regression models.

## Discussion

The main findings in this study are that (1) four favourable adiposity genetic variants are significantly associated with physical activity level, (2) favourable adiposity genetic liability protects against stage 2 hypertension depending on the individual’s BMI (3) in individuals with high BMI, physical activity provides additional protection against stage 2 hypertension independent of genetic liability. Our study provided insight into the complex relationship between favourable adiposity, BMI, physical activity, and the development of hypertension.

We identified four SNPs associated with physical activity none of which were implicated in physical activity previously. The exact mechanism by which these SNPs potentially enhance physical activity remains to be elucidated. However, for one SNP (rs4684847), the nearest gene is peroxisome proliferator-activated receptor gamma (*PPARG*) which influences muscle metabolism and insulin sensitivity ^37^, which are crucial for physical activity and exercise performance ^38,39^ implying a potential path to physical activity for this SNP. Three of these physical activity implicated SNPs (rs987469, rs4684847, rs12369179) are associated with BMI and the same allele that is associated with enhanced physical activity is also associated with a lower BMI implying that these SNPs may increase BMI by reducing physical activity levels. Future research focusing on the relationship between physical activity, BMI and these SNPs could shed more light on the matter. These SNPs were excluded from our analyses as among high body mass subgroups we observed and association between genetic liability and physical activity that was attenuated after excluding these SNPs.

In the current study we replicated previous findings that favourable adiposity genetic liability protects against hypertension ^13,14^ and we additionally found that the combined effect of physical activity and high genetic liability, predominantly in individuals with high BMI, enhances the protection against hypertension. High BMI groups of individuals carry up to 16% less risk of hypertension when carrying optimal status of both genetic and physical activity compared to when both genetic and physical activity are suboptimal. A person with high BMI could reduce risk of hypertension by 6-9% by engaging in physical activity alone when genetic liability status is kept constant. In our previous work ^11^ focusing on adiposity genetic liability, we observed a 10% reduction in risk of hypertension as a result of physical activity. However, our previous work did not differentiate the high and low body mass groups. The 6-9% risk reduction we observed as a result of physical activity in the current study is personalised depending on BMI and genetic liability status. This reduction of risk of hypertension is not observed in the low body mass group. Our previous work also demonstrated that a combination of suboptimal obesity genetic liability and suboptimal physical activity enhances risk of hypertension ^11^. However, our previous work did not differentiate between specific obesity subgroups, and it was focused on obesity genetic liability rather than favourable adiposity genetic liability (one is known to increase risk of hypertension and the other reduces risk of hypertension) ^11^. The current research additionally examines how favourable adiposity genetic liability reduces hypertension stratified by BMI and physical activity.

Our study is a step forward in personalised medicine compared to previous community intervention programs such as the “Pas-a-Pas” community intervention that observed a 6.63 mmHg reduction in SBP ^40^ as a result of physical activity.

Reduction in hypertension risk could alleviate the burden on the National Health Service (NHS), by decreasing hospital admissions, the need for antihypertensive medications ^41^, and the incidence of hypertension-related diseases such as stroke ^42^. Obesity, a major risk factor for hypertension, creates a substantial burden to the NHS, with obesity-related conditions costing the NHS £6.5 billion annually ^43^. Our study shows that physical activity interventions would only be beneficial in reducing risk of hypertension for high body mass groups suggesting that physical activity programs that aim to reduce hypertension should target high body mass groups essentially as there is no benefit for low body mass groups. Our study paves the way for precision medicine and improved classification of individuals at risk. The findings in our study provide potential impact on better management and prevention of hypertension among high body mass individuals. The results could guide health professionals to encourage high body mass individuals to engage in physical activity. By targeting high body mass groups, physical activity programs could mitigate the costs, improve patient outcomes, and enhance public health.

The protective effect of genetic liability on hypertension does not exist among individuals with low BMI no matter what their physical activity level is. One potential explanation for this could be that BMI reflects both muscle mass and fat mass ^44^. It is possible that the individuals we stratified in the low BMI group could have low muscle mass as well as low fat mass. Low muscle mass could be an indicator of subclinical underlying issues with potential impact on hypertension (e.g. sarcopenia ^45^ or chronic obstructive pulmonary disease (COPD) ^46, 47,48^). We have adjusted for factors such as smoking (strong risk factors for COPD) as well as age, and diet to mitigate the effect of such confounders, however, unknown confounding effects might still exist.

The main strengths of this study include the UKB’s large sample size and rich phenotyping, which provide optimal statistical power to investigate the relationship between genetic factors and complex outcomes. Another strength is the number of favourable adiposity variants used in this study utilising the most recent insight into capturing more accurate estimation of favourable adiposity genetic liability. Another strength is in the use of K-means algorithm ensuring that the self-reported ancestry data matches with the genetically derived ancestry and thus providing improved accuracy into the use of the European ancestry sample of the UKB.

While our findings are promising, further research is needed to explore the underlying mechanisms. Future studies should also investigate whether similar patterns are observed in different populations and age groups, and how these findings could be translated into practical interventions for hypertension prevention.

## Conclusion

The findings from this study demonstrate that the protective effect of favourable adiposity varies depending on the body mass and physical activity status. The finding underscores the significant role of physical activity in mitigating hypertension risk, particularly in individuals with high body mass and favourable adiposity genetic liability. Further research utilising ancestry-specific genetic variants could help understanding how these results apply to other ethnic populations.

## Author contributions

Conceptualization, R.P.; Data curation, C.H.; Formal analysis, C.H.; Funding acquisition, R.P.; Investigation, C.H.; Methodology, C.H. and R.P.; Project administration, R.P.; Resources, R.P.; Supervision, R.P.; Writing—original draft, C.H.; Writing—review and editing, C.H. and R.P. All authors have read and agreed to the published version of the manuscript.

## Funding and UKB application

UKB genotyping was supported by the British Heart Foundation (grant SP/13/2/30111) for Large-scale comprehensive genotyping of UKB for cardiometabolic traits and diseases: UK CardioMetabolic Consortium.

## Data Availability

Not applicable

## Acknowledgements

This research has been conducted using the UK Biobank Resource under Application Number 60549.

## Informed consent

Informed consent was obtained from all subjects involved in the study.

## Data availability statement

Not applicable.

## Institutional review board statement

The study was conducted in accordance with the Declaration of Helsinki and approved by the Institutional Review Board (or Ethics Committee) of Brunel University London, College of Health, Medicine and Life Sciences (27684-LR-Jan/2021–29 901-1).

## References

1. WHO. Global report on hypertension: the race against a silent killer. Geneva: World Health Organization; 2023. Licence: CC BY-NC-SA 3.0 IGO. Https://WwwWhoInt/Publications/i/Item/9789240081062 2023:1–291.

2. de Oliveira CM, Ulbrich AZ, Neves FS, Dias FAL, Horimoto ARVR, Krieger JE, et al. Association between anthropometric indicators of adiposity and hypertension in a Brazilian population: Baependi Heart Study. PLoS One 2017;12:e0185225. doi:10.1371/journal.pone.0185225.

3. Khaleghi MM, Jamshidi A, Afrashteh S, Emamat H, Farhadi A, Nabipour I, et al. The association of body composition and fat distribution with hypertension in community-dwelling older adults: the Bushehr Elderly Health (BEH) program. BMC Public Health 2023;23:1–11. doi:10.1186/S12889-023-16950-8/TABLES/3.

4. Guo X, Xu Y, He H, Cai H, Zhang J, Li Y, et al. Visceral fat reduction is positively associated with blood pressure reduction in overweight or obese males but not females: an observational study. Nutr Metab (Lond) 2019;16:44. doi:10.1186/s12986-019-0369-0.

5. Hwang Y-C, Fujimoto WY, Kahn SE, Leonetti DL, Boyko EJ. Greater visceral abdominal fat is associated with a lower probability of conversion of prehypertension to normotension. J Hypertens 2017;35:1213–1218. doi:10.1097/HJH.0000000000001296.

6. Wang Z, Zeng X, Chen Z, Wang X, Zhang L, Zhu M, et al. Association of visceral and total body fat with hypertension and prehypertension in a middle-aged Chinese population. J Hypertens 2015;33:1555–1562. doi:10.1097/HJH.0000000000000602.

7. Locke A, Kahali B, Speliotes E. Genetic studies of body mass index yield new insights for obesity biology. Nature 2015;518:197–206. doi:10.1038/nature14177.

8. Yengo L, Sidorenko J, Kemper KE, Zheng Z, Wood AR, Weedon MN, et al. Meta-analysis of genome-wide association studies for height and body mass index in ∼700000 individuals of European ancestry. Hum Mol Genet 2018;27:3641–3649. doi:10.1093/hmg/ddy271.

9. Shungin D, Winkler T, Croteau-Chonka DC, Ferreira T, Locke AE, Mägi R, et al. New genetic loci link adipose and insulin biology to body fat distribution. Nature 2015 518:7538 2015;518:187–196. doi:10.1038/nature14132.

10. Giontella A, Lotta LA, Overton JD, Baras A, Minuz P, Melander O, et al. Causal Effect of Adiposity Measures on Blood Pressure Traits in 2 Urban Swedish Cohorts: A Mendelian Randomization Study. Journal of the American Heart Association: Cardiovascular and Cerebrovascular Disease 2021;10:20405. doi:10.1161/JAHA.120.020405.

11. Hezekiah C, Blakemore AI, Bailey DP, Pazoki R. Physical activity alters the effect of genetic determinants of adiposity on hypertension among individuals of European ancestry in the UKB. Scand J Med Sci Sports 2024;34:e14636. doi:10.1111/SMS.14636.

12. Martin S, Cule M, Basty N, Tyrrell J, Beaumont RN, Wood AR, et al. Genetic Evidence for Different Adiposity Phenotypes and Their Opposing Influences on Ectopic Fat and Risk of Cardiometabolic Disease. Diabetes 2021;70:1843– 1856. doi:10.2337/db21-0129.

13. Ahmed A, Justo S, Yaghootkar H, Hanieh Yaghootkar C, Banks J. Genetic scores associated with favourable and unfavourable adiposity have consistent effect on metabolic profile and disease risk across diverse ethnic groups. Diabetic Medicine 2023;40:e15213. doi:10.1111/DME.15213.

14. Yaghootkar H, Lotta LA, Tyrrell J, Smit RAJ, Jones SE, Donnelly L, et al. Genetic Evidence for a Link Between Favorable Adiposity and Lower Risk of Type 2 Diabetes, Hypertension, and Heart Disease. Diabetes 2016;65:2448– 2460. doi:10.2337/DB15-1671.

15. Jackson C, Herber-Gast GC, Brown W. Joint Effects of Physical Activity and BMI on Risk of Hypertension in Women: A Longitudinal Study. J Obes 2014;2014. doi:10.1155/2014/271532.

16. UK Biobank. UK Biobank: Protocol for a large-scale prospective epidemiological resource (AMENDMENT ONE FINAL). 2007.

17. Elliott P, Peakman TC. The UK Biobank sample handling and storage protocol for the collection, processing and archiving of human blood and urine. Int J Epidemiol 2008;37:234–244. doi:10.1093/ije/dym276.

18. General Assembly of the World Medical Association. World Medical Association Declaration of Helsinki: ethical principles for medical research involving human subjects. J Am Coll Dent 2014;81:14–18.

19. Bycroft C, Freeman C, Petkova D, Band G, Elliott LT, Sharp K, et al. Genome-wide genetic data on ∼500,000 UK Biobank participants. BioRxiv 2017:166298. doi:10.1101/166298.

20. Marchini J, O’Connell J, Delaneau ,, Jonathan, Sharp Kevin, Kretzschmar Warren, Band G, et al. UK Biobank Phasing and Imputation Documentation Contributors to UK Biobank Phasing and Imputation. Https://WwwJiscmailAcUk/Cgi-Bin/Webadmin?A0=UKB-GENETICS 2015.

21. Welsh S, Peakman T, Sheard S, Almond R. Comparison of DNA quantification methodology used in the DNA extraction protocol for the UK Biobank cohort. BMC Genomics 2017;18. doi:10.1186/S12864-016-3391-X.

22. Tobin MD, Sheehan NA, Scurrah KJ, Burton PR. Adjusting for treatment effects in studies of quantitative traits: antihypertensive therapy and systolic blood pressure. Stat Med 2005;24:2911–2935. doi:10.1002/SIM.2165.

23. Whelton PK, Carey RM, Aronow WS, Casey DE, Collins KJ, Dennison Himmelfarb C, et al. 2017 ACC/AHA/AAPA/ABC/ACPM/AGS/APhA/ASH/ASPC/NMA/PCNA Guideline for the Prevention, Detection, Evaluation, and Management of High Blood Pressure in Adults: Executive Summary: A Report of the American College of Cardiology/American Heart Association Task Force on Clinical Practice Guidelines. J Am Coll Cardiol 2018;71:2199–2269. doi:10.1016/j.jacc.2017.11.005.

24. Craig CL, Marshall AL, Sjöström M, Bauman AE, Booth ML, Ainsworth BE, et al. International physical activity questionnaire: 12-country reliability and validity. Med Sci Sports Exerc 2003;35:1381–1395. doi:10.1249/01.MSS.0000078924.61453.FB.

25. Cassidy S, Chau JY, Catt M, Bauman A, Trenell MI. Cross-sectional study of diet, physical activity, television viewing and sleep duration in 233 110 adults from the UK Biobank; the behavioural phenotype of cardiovascular disease and type 2 diabetes. BMJ Open 2016;6:e010038. doi:10.1136/bmjopen-2015-010038.

26. IPAQ. Guidelines for Data Processing and Analysis of the International Physical Activity Questionnaire (IPAQ)-Short and Long Forms. 2005.

27. WHO Consultation on Obesity (1999: Geneva S, World Health Organization. WHO Technical Report Series OBESITY: PREVENTING AND MANAGING THE GLOBAL EPIDEMIC 2000.

28. UK Biobank. Showcase: Data-Field 20161 2016. https://biobank.ndph.ox.ac.uk/showcase/field.cgi?id=20161 (accessed June 29, 2023).

29. UK Biobank. Showcase: Data-Field 20117 2016. https://biobank.ndph.ox.ac.uk/showcase/field.cgi?id=20117 (accessed February 28, 2024).

30. UK Biobank. UK Biobank Biomarker assay quality procedures: approaches used to minimise systematic and random errors (and the wider epidemiological implications). Http://WwwUkbiobankAcUk/ 2019.

31. Machiela MJ, Chanock SJ. LDlink: a web-based application for exploring population-specific haplotype structure and linking correlated alleles of possible functional variants. Bioinformatics 2015;31:3555–3557. doi:10.1093/bioinformatics/btv402.

32. Chang CC, Chow CC, Tellier LCAM, Vattikuti S, Purcell SM, Lee JJ. Second-generation PLINK: Rising to the challenge of larger and richer datasets. Gigascience 2015;4:7. doi:10.1186/S13742-015-0047-8/2707533.

33. Purcell S, Chang C. PLINK 2.0 2016. https://www.cog-genomics.org/plink/2.0/ (accessed August 10, 2024).

34. MacQueen J. Some methods for classification and analysis of multivariate observations. Fifth Berkeley Symposium on Mathematical Statistics and Probability, Berkeley: University of California Press; 1967, p. 281–297.

35. Henry D, Dymnicki AB, Mohatt N, Allen J, Kelly JG. Clustering Methods with Qualitative Data: A Mixed Methods Approach for Prevention Research with Small Samples. Prev Sci 2015;16:1007. doi:10.1007/S11121-015-0561-Z.

36. R Core Team. R: The R Project for Statistical Computing 2021. https://www.r-project.org/ (accessed September 23, 2021).

37. Poulsen P, Andersen G, Fenger M, Hansen T, Echwald SM, Vølund A, et al. Impact of Two Common Polymorphisms in the PPARγ Gene on Glucose Tolerance and Plasma Insulin Profiles in Monozygotic and Dizygotic TwinsThrifty Genotype, Thrifty Phenotype, or Both? Diabetes 2003;52:194–198. doi:10.2337/DIABETES.52.1.194.

38. Prior SJ, Blumenthal JB, Katzel LI, Goldberg AP, Ryan AS. Increased Skeletal Muscle Capillarization After Aerobic Exercise Training and Weight Loss Improves Insulin Sensitivity in Adults With IGT. Diabetes Care 2014;37:1469– 1475. doi:10.2337/dc13-2358.

39. Shepherd SO, Wilson OJ, Taylor AS, Thøgersen-Ntoumani C, Adlan AM, Wagenmakers AJM, et al. Low-Volume High-Intensity Interval Training in a Gym Setting Improves Cardio-Metabolic and Psychological Health. PLoS One 2015;10:e0139056. doi:10.1371/journal.pone.0139056.

40. Arija V, Villalobos F, Pedret R, Vinuesa A, Jovani D, Pascual G, et al. Physical activity, cardiovascular health, quality of life and blood pressure control in hypertensive subjects: randomized clinical trial. Health Qual Life Outcomes 2018;16. doi:10.1186/S12955-018-1008-6.

41. di Cagno A, Fiorilli G, Buonsenso A, Di Martino G, Centorbi M, Angiolillo A, et al. Long-Term Physical Activity Effectively Reduces the Consumption of Antihypertensive Drugs: A Randomized Controlled Trial. Journal of Cardiovascular Development and Disease 2023, Vol 10, Page 285 2023;10:285. doi:10.3390/JCDD10070285.

42. Hooker SP, Diaz KM, Blair SN, Colabianchi N, Hutto B, McDonnell MN, et al. Association of Accelerometer-Measured Sedentary Time and Physical Activity With Risk of Stroke Among US Adults. JAMA Netw Open 2022;5:e2215385– e2215385. doi:10.1001/JAMANETWORKOPEN.2022.15385.

43. 43. Bell M, Deyes K. ESTIMATING THE FULL COSTS OF OBESITY frontier economics A report for Novo Nordisk. Https://WwwFrontier-EconomicsCom/Media/Hgwd4e4a/the-Full-Cost-of-Obesity-in-the-UkPdf 2022. https://www.frontier-economics.com/media/hgwd4e4a/the-full-cost-of-obesity-in-the-uk.pdf (accessed October 6, 2024).

44. Romero-Corral A, Somers VK, Sierra-Johnson J, Thomas RJ, Collazo-Clavell ML, Korinek J, et al. Accuracy of body mass index in diagnosing obesity in the adult general population. Int J Obes 2008;32:959–966. doi:10.1038/ijo.2008.11.

45. Curtis M, Swan L, Fox R, Warters A, O’Sullivan M. Associations between Body Mass Index and Probable Sarcopenia in Community-Dwelling Older Adults. Nutrients 2023, Vol 15, Page 1505 2023;15:1505. doi:10.3390/NU15061505.

46. Attaway AH, Lopez R, Welch N, Bellar A, Hatipoğlu U, Zein J, et al. Muscle loss phenotype in COPD is associated with adverse outcomes in the UK Biobank. BMC Pulm Med 2024;24:1–11. doi:10.1186/S12890-024-02999-7/FIGURES/2.

47. Han K, Park YM, Kwon HS, Ko SH, Lee SH, Yim HW, et al. Sarcopenia as a Determinant of Blood Pressure in Older Koreans: Findings from the Korea National Health and Nutrition Examination Surveys (KNHANES) 2008–2010. PLoS One 2014;9:e86902. doi:10.1371/JOURNAL.PONE.0086902.

48. Kim SH, Park JH, Lee JK, Heo EY, Kim DK, Chung HS. Chronic obstructive pulmonary disease is independently associated with hypertension in men. Medicine (United States) 2017;96. doi:10.1097/MD.0000000000006826.

